# Impact of a Chlorhexidine Decolonization on the Nasal and Dermal Microbiome of Therapy Dogs Participating in Hospital Animal-Assisted Intervention Programs: A Pilot Study

**DOI:** 10.1101/2021.02.11.21250783

**Authors:** Kathryn R. Dalton, Kathy Ruble, Karen C. Carroll, Laurel E. Redding, Allen R. Chen, Elizabeth A. Grice, Daniel O. Morris, Meghan F. Davis

## Abstract

Therapy animals in hospital animal-assisted intervention programs are an invaluable part of holistic patient care. However, therapy dogs may be exposed to hospital-associated pathogens through these activities. This pilot study sought to examine the effect of topical chlorhexidine application, used as an infection control measure, on the microbial composition of the skin and mucous membranes of therapy dogs. We found that the chlorhexidine decolonization intervention altered microbial alpha diversity and shifted microbial structures in these therapy dogs, and particularly influenced more phylogenetically rare taxa. Specifically, the intervention reduced the abundance of *Staphylococcus pseudintermedius*, the archetypal canine commensal and a frequent cause of opportunistic infections. However, it did not reduce levels of *S. aureus*, which is a common hospital-associated pathogen of people. These preliminary findings highlight the importance of considering holistic microbial communities when undertaking infection control strategies, and stress the need for further research to understand the unintended consequences of antiseptic use on therapy dogs.

## Introduction

The benefits of the human-animal bond have extended into the use of animal-assisted interventions, which are increasingly used in healthcare facilities for their widely recognized benefits (Kamioka et al., 2014; Waite et al., 2018). However, risks to both human patients and therapy animals from exposure to hospital-associated pathogens are not fully characterized. Previous work has shown that therapy animals which volunteer in hospitals are five times more likely to carry methicillin-resistant *Staphylococcus aureus* (MRSA) compared to therapy animals that volunteer in non-healthcare settings (Lefebvre et al., 2009). Thus, infection control strategies designed to reduce the spread of pathogens between patients and therapy animals are needed to improve program safety and increase their beneficial utilization.

A common infection control practice is the use of disinfectants on frequently touched environmental surfaces, or fomites, including the fur and gear of therapy animals. Topical antiseptics have previously been shown to reduce the bacterial burden of *Staphylococcus aureus* on therapy animals, as well as reduce the transmission to patients, using culture-based methods (Dalton et al., 2018). However, antiseptics have been shown to alter the microbial composition of both humans and dogs. In humans, the effects of topical antiseptics were dependent on individual and body site-specific colonization signatures but lowered the overall microbial diversity level (SanMiguel et al., 2018). In dogs, chlorhexidine antiseptics are frequently used to treat atopic dermatitis, which is characterized by a higher abundance of staphylococcal species and lower overall microbial diversity (Chermprapai et al., 2019; Tress et al., 2017). Treatment with a topical antiseptic and targeted systemic antimicrobial therapy for staphylococcal pyoderma restored the dermal microbiome in atopic patients (Bradley et al., 2016). It is unclear what impact topical antiseptics might have on the skin and nasal microbial communities of healthy therapy animals. Therapy animals, with their frequent hospital exposure, may have unique microbial compositions compared to typical pet dogs. These distinct communities could then be differentially affected by chlorhexidine or similar disinfectant procedures.

The goal of this research was to describe the skin and nasal microbial composition of therapy dogs and determine how these microbial communities are influenced by the use of a chlorhexidine topical intervention. The results of this work will inform the design of more extensive studies, with implications for infection control strategies. Such work is critical to minimize unintended consequences to the health of these volunteer therapy animals from interventions such as the use of antiseptics.

## Methods

### Study Population

This pilot study was conducted in a mid-Atlantic hospital between July 2016 and May 2017. The study protocol was approved by all applicable institutional review boards, institutional animal care and use committees, and scientific review committees prior to data collection. Canine participants were registered therapy animals volunteering in an academic hospital’s animal-assisted intervention (AAI) program. The therapy visits were group sessions, lasting one hour, where multiple pediatric patients interacted with the animals in a large multi-use space within a children’s hospital.

### Data Collection

Enrolled therapy dogs underwent two observational control visits, where they adhered to existing hospital policies, requiring the dog to be bathed using an over-the-counter shampoo of the owner’s choice within 24 hours prior to entering the hospital. Matched samples were collected from all individuals before and after the therapy session. All sample collection was done by trained research staff.

A detailed description of sample collection procedure is described elsewhere (Dalton et al., 2021). Briefly, we obtained nasal, oral, inguinal, and perineal samples from the therapy dog before and after every therapy session. Samples were collected using sterile flocked swabs (Puritan, Guilford, ME, USA), then stored at −80°C until processing. Handlers were asked questions about their therapy dog’s medical history and volunteer work at each visit. We captured the total number of patients who interacted with the dog at every visit.

### Intervention

After two control visits, the therapy dog team underwent two intervention visits using a two-part decolonization regimen on the therapy dog. Prior to the visit, therapy dog handlers bathed their dogs with a 4% chlorhexidine-based veterinary prescription shampoo (DUOXO Ceva, Libourne, France), as per the manufacturer’s directions. During the therapy visits, the dogs were wiped with pre-moistened cloths containing 3% chlorhexidine gluconate, 0.5% climbazole and 0.05% phytosphingosine salicyloyl (DOUXO 3% chlorhexidine pads PS; Ceva, Libourne, France) across the dorsal head and back (i.e., the “petting zone”). Handlers were given information about both products before usage. The same data collection protocol was implemented for these intervention visits, as described above.

### Laboratory Processing for Microbial Communities

Detailed descriptions of laboratory and sequencing protocols are available elsewhere (Dalton et al., 2021). Briefly, DNA was extracted from the thawed sterile flocked swabs, and the V1-3 region of the 16S rRNA gene was PCR-amplified and sequenced using established protocols for microbial composition analysis using the Illumina MiSeq (Illumina, San Diego, CA) at the University of Pennsylvania Next Generation Sequencing Core (Fadrosh et al., 2014).

The QIIME (v2.7) pipeline was used to match sequencing reads to samples (Bolyen et al., 2019), and the DADA2 pipeline was used for quality filtering and clustering samples into features (amplicon sequence variants, ASVs) (Callahan et al., 2016). ASVs were matched to taxonomy and phylogeny using established pipelines (Dalton et al., 2021). Unidentified sequences not matched to taxonomy on our classifier were manually entered into the NCBI BLAST database for taxonomic classification. Quality control was ensured through identifying likely taxa contaminants through negative controls collected during field sampling, DNA extraction, and sequencing (Davis et al., 2018).

### Statistical Analysis

Statistical analyses were performed in RStudio v1.1.423 (R Development Core Team, 2010). To maintain the maximum number of samples for comparison, the sequencing data were not rarefied for statistical analysis. Taxa tables, and matching phylogeny and taxonomy, were analyzed using the *phyloseq* pipeline to calculate alpha (within sample) and beta (between samples) diversity metrics (McMurdie & Holmes, 2013). The negative binomial-based model DESeq2 program was used to identify differential abundance of key taxa between groups (Love et al., 2014). Kruskal-Wallis nonparametric one-way analysis of variance test was used to examine differential alpha diversity between all groups, and Wilcoxon rank-sum test was used for pair-wise comparisons between groups, both adjusted for multiple comparisons using the Benjamini-Hochberg false discovery rate (FDR) correction.

## Results

### Study Population and Samples

A total of 4 dogs were included in the study over 13 visits, with 5 intervention study visits, shown in **Table 1**. Ages ranged from 18 to 132 months old, and more dogs (n =3) were female.

**Table 1:** Study Population.

|  | **Total** |
| --- | --- |
| **Unique Dogs** | 4 |
| Age < 6 years old | 2 (50%) |
| Male | 1 (25%) |
| **Study Visits** | 13 |
| Intervention Visits | 5 (38.5%) |
| Antibiotics Last Month | 5 (38.5%) |
| Veterinary Hospital Visit Last Month | 3 (23%) |
| AAI Frequency > once per week | 5 (38.5%) |
| Patients in Visits Total > 3 | 7 (54%) |

Therapy dog handlers reported recent veterinary antimicrobial usage (either topical or systemic) within the last month at 38% of the visits, and a recent veterinary hospital visit in the last month at 23% of the visits. Therapy dog handlers reported being involved in AAI programs within the last week at 38% of the visits. Of the 13 visits, 54% had more than 3 patients interacting with the dog during the therapy visit. A total of 100 swabs were collected for microbial analysis - 26 each nasal, oral and perineal swabs, and 22 inguinal samples. All samples were matched for pre- and post-visits.

### Taxa Abundance

The relative abundance of taxa was different both across sample locations and individual dogs, as shown in **Figure 1**.**A-D**. Both the nasal and oral samples tended to be dominated by key taxa, such as *Staphylococcus* (mean relative abundance 0.111), *Porphyromonas* (mean 0.099), *Corynebacterium* (mean 0.095), and *Moraxella* (mean 0.073) genera, while inguinal samples had a higher number of more unique taxa.

**Figure 1:**
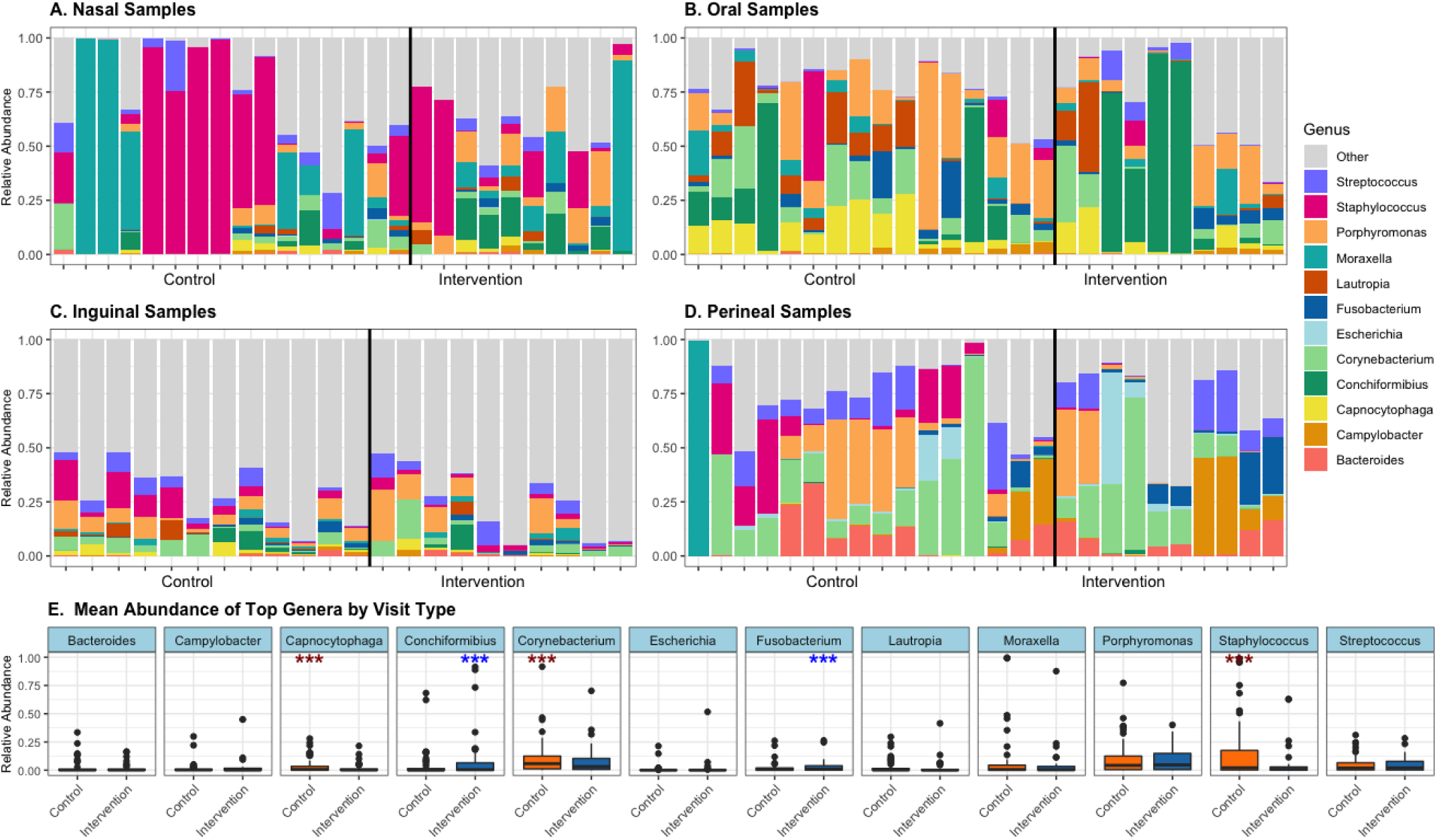
Relative Abundance of Most Abundant Genera, by Site and Visit Type*(genera with mean abundance >25%)* A-D: Bars represent individual sample-level relative abundance of genera that have a mean abundance above 25%, stratified by site. Black horizontal bar divides samples taken from control visits and intervention visits. E: Mean relative abundance of top genera by visit type, aggregated pre/post samples and different sites. *** DESeq results p<0.0001 for differential abundance by visit type, red = higher in control, blue = higher in intervention

The intervention impacted levels of the most abundant genera in different ways, as shown in **Figure 1**.**E**. The intervention reduced the mean relative abundance of *Staphylococcus, Corynebacterium*, and *Capnocytophaga* taxa (DESeq p<0.0001). Conversely, *Conchiformibius* and *Fusobacterium* genera were more abundant following intervention visits (DESeq p<0.0001). No significant differences were found in these taxa between pre-and post-visit samples overall, and within each site.

### Staphylococcal Species Abundance

**Figure 2** shows the relative abundance of the three most abundant *Staphylococcus* species - *S. aureus, pseudintermedius*, and *schleiferi* (absolute abundance depicted in **Supplemental Figure 1**). No significant changes occurred between pre and post samples. However, there was a significant difference in the relative abundance of *S. pseudintermedius* between control and intervention samples among pre-visit samples (relative abundance 0.043 in control and 0.0008 intervention, Wilcoxon rank-sum test p=0.002) and post-visit samples (relative abundance 0.042 in control and 0.004 intervention, Wilcoxon rank-sum test p=0.008).

**Figure 2:**
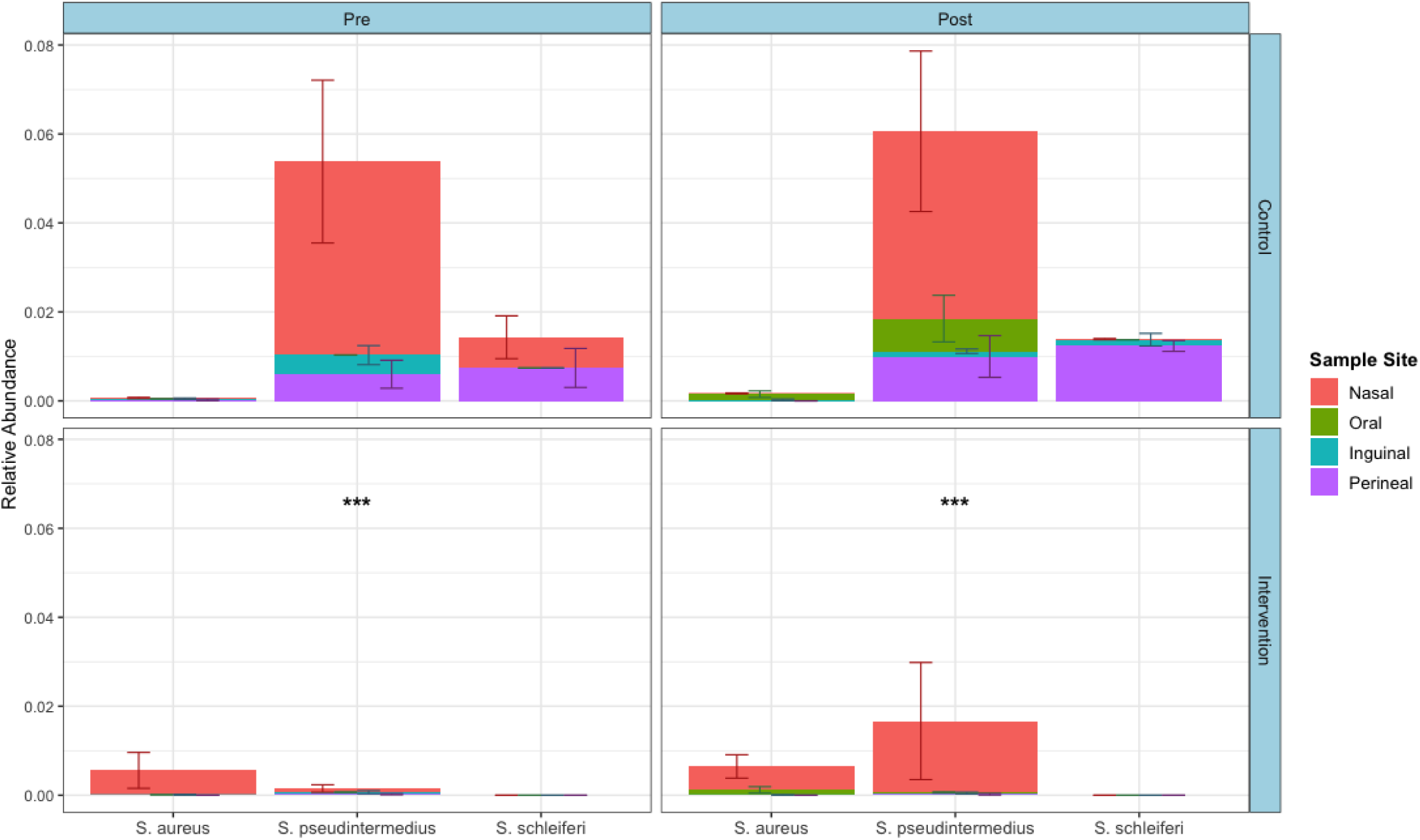
Relative Abundance of Key Staphylococcal Species, by Visit Type and Time. Bars are aggregated mean relative abundance of top three *Staphylococcus* species, colored by sample site, with error bars for standard error. *** Wilcoxon Rank-Sum Test continuity-corrected p<0.01 Control vs Intervention.

*S. schleiferi* abundance also decreased following intervention visits (mean relative abundance 0.026 in control and 0 in intervention, Wilcoxon p=0.036), seen in both pre and post samples. *S. aureus* levels were not significantly different between control and intervention visits for either pre-visit or post-visit samples.

### Alpha Diversity

Alpha diversity by site was evaluated with Shannon and Faith’s phylogenetic metrics **(Figure 3**.**A&B)**. The alpha diversity of nasal, oral and perineal samples were similar to each other and significantly different from that of inguinal samples (Kruskal-Wallis test p<0.001 for all pairwise comparisons with inguinal samples, for Shannon and Faith’s). According to the Wilcoxon rank-sum test, there was no statistical difference in the overall alpha diversity levels between pre- to post-visit overall, within each site, and across individual dogs.

**Figure 3:**
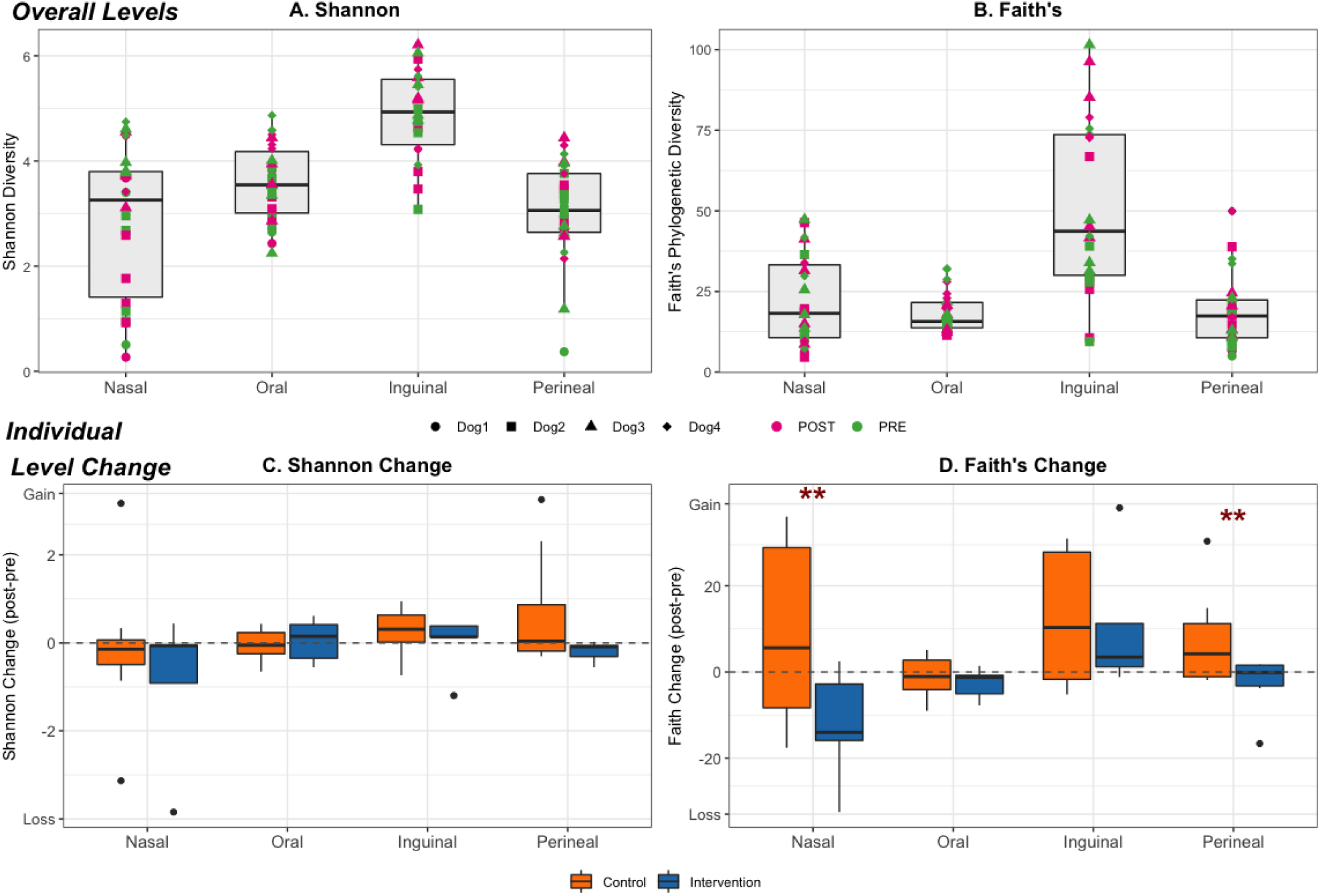
Shannon and Faith’s Alpha Diversity Levels by Sample Site *(overall and changes during visit)* A & B aggregated alpha diversity levels (Shannon A, and Faith B), by sample site, C & D individual level change (post-pre visit) alpha diversity levels by sample site *** Wilcoxon-Test p<0.05”, “Control vs Intervention

At the individual dog level **(Figure 3**.**C&D**), there was an overall increase in alpha diversity after control visits (mean Shannon change 0.18, mean Faith change 6.54) and an overall decrease following intervention visits (mean Shannon change −0.26, mean Faith change −1.93), with the most pronounced differences occurring in perineal and nasal samples (significant difference in Faith metric using Wilcoxon rank-sum test p<0.05).

**Supplemental Table 1** shows how additional dog demographics such as age, sex, and medical history, impacted alpha diversity levels. Significant associations were observed based on recent antibiotic usage and veterinary hospital visit, as well as reported AAI frequency (the number of times the dog participated in AAI programs) and the number of patients in the visit.

### Beta Diversity

Principal coordinate analysis plots are displayed in **Figure 4**, overall and within each site, using both weighted UniFrac and unweighted UniFrac distance. Clustering was observed by sample site (PERMANOVA FDR-corrected p<0.001 for both weighted and unweighted UniFrac distance) and by subject overall and within each individual site (PERMANOVA p<0.001 for both weighted and unweighted UniFrac distance). There were no differences in microbial composition when comparing before versus after the visit, overall and within each site.

**Figure 4:**
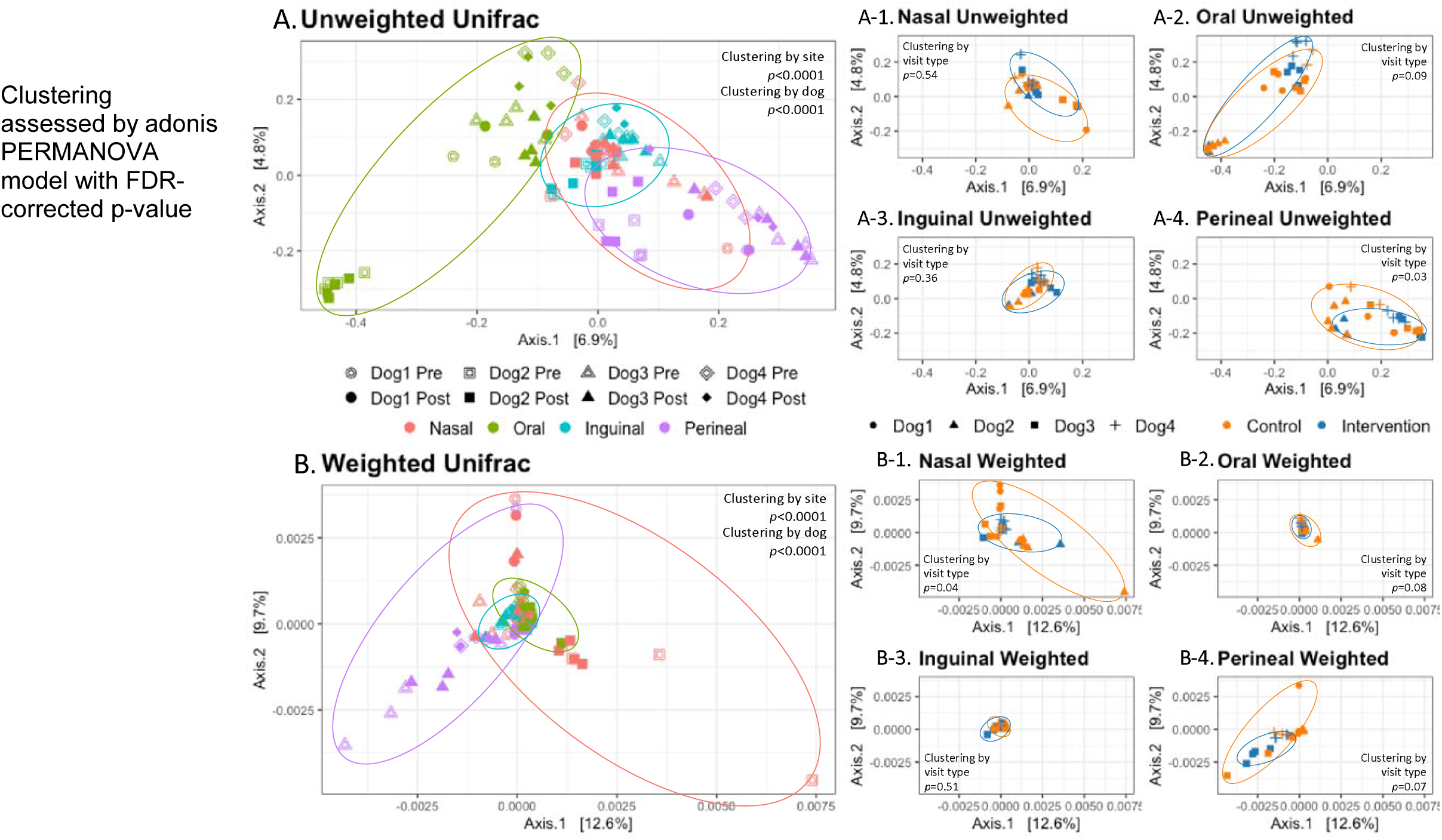
Principal Coordinate Plots for Unweighted and Weighted UniFrac Beta Diversity Levels by Dog and Site, and by Visit Type within Each Site.

There was a difference in microbial composition between samples from control visits and samples from intervention visits (PERMANOVA p=0.03 unweighted and p=0.009 weighted). The impact of the intervention on microbial composition was different based on the sample site. When using unweighted UniFrac metrics, only perineal samples showed microbial composition differences between control and intervention visits (PERMANOVA p=0.03). Using the weighted UniFrac metric, only nasal samples were significantly different by PERMANOVA (p-value below 0.05). There was no significant difference in intervention effect when stratifying by pre-visit or post-visit.

**Supplemental Table 1** shows the distance between two samples by dog demographic factors, such as age, sex, and medical history. There was a significant difference in the microbial composition of dogs based on their age, sex, and recent antibiotic usage, using both unweighted and weighted UniFrac distance. Recent veterinary hospital exposure and the number of patients in the therapy visit were shown to impact the unweighted but not weighted UniFrac distance, while reported AAI frequency did not have an association.

## Discussion

This research aimed to explore the microbiota of healthy, non-atopic dogs that participate in an AAI program and to examine the influence of topical chlorhexidine use on therapy dogs in the context of a pilot infection control intervention. We found that body sites were uniquely affected by the chlorhexidine intervention, and more phylogenetically rare taxa and canine-specific taxa – such as *S. pseudintermedius* and *S. schleiferi* – were less abundant following the intervention.

### Therapy Dogs Compared to Typical Pet Dogs

As in typical companion dogs, therapy dogs showed unique microbial patterns across body sites. Samples were more similar to the same site on another individual dog than to another site on the same individual, confirming that the ecological body site niche is a more significant determinant of microbiota composition (Chermprapai et al., 2019; Cuscó et al., 2017; Grice et al., 2009; Misic et al., 2015). As with humans, the skin microbiota in our canine participants varied between different body sites, presumably because of differences in the local cutaneous microclimate. Previous studies have shown that the canine bacterial community is diverse and variable across different body sites within the same dog, and across the same site in different dogs (Hoffmann et al., 2014).

Alpha diversity metrics, assessed by Shannon’s and Faith’s Phylogenetic diversity, were shown to be unique across body sites, with nasal samples dominated by few key taxa and inguinal samples having a higher number of more unique taxa. As was seen in previous studies, therapy dogs had higher diversity levels in haired regions (inguinal skin) than mucosal areas and mucocutaneous junctions (Hoffmann et al., 2014). *Staphylococcus* abundance was increased in these therapy dogs, particularly in nasal samples, in contrast to prior reports where *Moraxellla* tended to be the dominant taxa (Bradley et al., 2016; Tress et al., 2017). Nonetheless, *Staphylococcus* spp. levels in these therapy dogs were lower than what is reported in dogs with underlying pathologies, such as atopic dermatitis, pyoderma, or chronic rhinosinusitis (Bradley et al., 2016; Tress et al., 2017; Weese, 2013). Although no comparison dogs were sampled for this pilot study, the differences between the findings from this study and those reported in previous literature may suggest that frequent hospital exposures could affect their microbial composition of these dogs. This same finding has been reported in healthcare workers when comparing their microbial composition to the general population (Rosenthal et al., 2013).

### Effect of the Decolonization Intervention on Microbiota

Diversity metrics, both alpha (within-sample) and beta (between-sample), were influenced by the chlorhexidine in similar ways. Alpha diversity of samples tended to increase after control visits and decrease after intervention visits. There were minimal changes in alpha levels and beta microbial composition within samples following visits. However, there was a significant difference between control and intervention samples. Intervention samples tended to have lower alpha diversity and significantly different microbial compositions compared to control samples. This association was more robust when using phylogenetically weighted metrics (Faith’s alpha diversity and weighted UniFrac beta diversity) and within nasal and perineal sites.

Taking the results from both alpha and beta diversity together, the differences associated with the chlorhexidine intervention appear driven primarily by more phylogenetically rare taxa rather than common taxa. Previous studies on pet dogs (Davis, 2016; Oh et al., 2015; Song et al., 2013) have shown that pet dogs have more diverse unique microbial communities compared to their human counterparts. It can be proposed that these phylogenetically rare taxa originate from the dog and are reduced by application of chlorhexidine. Nasal and perineal sites, which both tend to have lower diversity levels overall, are more influenced by this disturbance. The finding that a topical treatment could be associated with altered microbiome on sites not directly exposed to the chlorhexidine (shampoo just on the skin, and wipes just on the dorsal back/head) is of great interest and unexpected.

To determine which microorganisms were associated with this change in diversity, we evaluated the abundance of identified taxa, which were differentially impacted by the chlorhexidine intervention. *Staphylococcus, Corynebacterium*, and *Capnocytophaga* species levels were shown to be reduced following the intervention, while *Fusobacterium* and *Conchiformibus* levels tended to be increased following the intervention. *Staphylococcus* and *Corynebacterium* spp. have been shown to be higher in abundance in dogs with medical conditions, such as atopic dermatitis (Hoffmann et al., 2014; Pierezan et al., 2016). In one study, *Capnocytophaga* was found in be increased in nasal neoplastic samples compared to healthy nasal samples (Tress et al., 2017). Studies on the effect of chlorhexidine on human skin have also shown decreases in overall diversity levels and decreases in specific *Staphylococcus* and *Corynebacterium* taxa (SanMiguel et al., 2018).

Due to its clinical significance, *Staphylococcus* was further evaluated at the species level. The chlorhexidine intervention impacted the abundance of *S. pseudintermedius*, and to a lesser degree *S. schleiferi*, more than *S. aureus. S. pseudintermedius* is the dominant coagulase-positive commensal of canine skin and mucous membranes, and the most common cause of canine skin and soft tissue infections. *S. schleiferi* is a less prevalent commensal of dogs, and most commonly causes skin and ear canal infections in the context of prior antimicrobial exposures. *S. aureus* is not considered to be part of the canine commensal microflora, and causes infection of dogs rarely (Morris et al., 2017). The abundance of both *S. pseudintermedius* and *S. schleiferi* decreased following the intervention, with this difference being significant for *S. pseudintermedius*. In contrast, *S. aureus* abundance increased slightly following intervention visits (in both relative and absolute levels). Again, even though the intervention was restricted to the skin/coat, nasal samples showed the greatest difference. This is not surprising because *Staphylococcus* species frequently colonize the nose in dogs (Iverson et al., 2015; Weese & van Duijkeren, 2010). *S. pseudintermedius* and *S. schleiferi* tend to be dog-specific microbiota and tend to only incidentally contaminant humans (Weese & van Duijkeren, 2010).

It is uncertain from our single timepoint samples if the intervention selectively removed or reduced the abundance of certain taxa, such as dog-specific *S. pseudintermedius, S. schleiferi*, and phylogenetically diverse microbiota. Another possibility is that the decolonization removed or reduced all microbiota equally, but prior to using the wipes and sampling, the therapy dog was recolonized with microbiota that are more commonly associated with humans or the hospital environment. This would result in what appeared to be no change in the abundance of common taxa, such as *S. aureus*. The source of this re-colonization could be from hospital exposure, from interaction with other individuals while going to the therapy visit (prior to our pre-visit sampling), or even from the therapy dog’s handler.

### Limitations and Future Directions

A limitation of this pilot study is that our samples only reflect carriage of microorganisms at one timepoint. We cannot make inferences from our data on whether these microbial exposures and observed changes are transient contamination or stable colonization. Future research in this field will examine the temporal progression and stability of microbial community alterations due to the decolonization; abundance of key taxa of clinical concern and dog-specific microbiota, as well as overall diversity levels. Further study should also aim to understand the secondary health consequences of the decolonization intervention within this therapy dog population, including the role of *S. pseudintermedius* as a protective commensal versus opportunistic pathogen, and possible competition with *S. aureus*. The zoonotic potential of both of these organisms is important to consider as these therapy dogs frequently interact with patients with compromised immune function.

## Conclusions

This study benefits from being the first to examine the microbiota of therapy dogs and the effect of a decolonization intervention on therapy animals in hospital AAI programs. Understanding cutaneous microbial ecology is essential to create future targeted therapies that might require not only a reduction in exposure to pathogenic bacteria, but also a promotion of the symbiotic commensal microbiota. This pilot study presents the feasibility and importance of assessing holistic microbial communities in this essential canine worker population. The study demonstrates the potential for infection control practices, designed to limit exposure to pathogens, to alter microbial communities more broadly, with unknown consequences. This has important implications for handlers and practitioners in charge of these therapy dogs’ care and hospital administration. Namely, the importance of considering the holistic microbial environment should be considered when designing interventions to keep hospital animal-assisted intervention programs safe for patients and therapy dogs.

## Data Availability

The raw sequence data and metadata can be found in NCBI Sequence Read Archive (SRA) database under BioProject PRJNA695069, BioSample SAM17600695. Unix and R code used for analysis can be found under KRD's Github repository: https://github.com/kathryndalton/AAT_pilot_analysis.

## Supplementary Materials

**Supplemental Figure 1:**
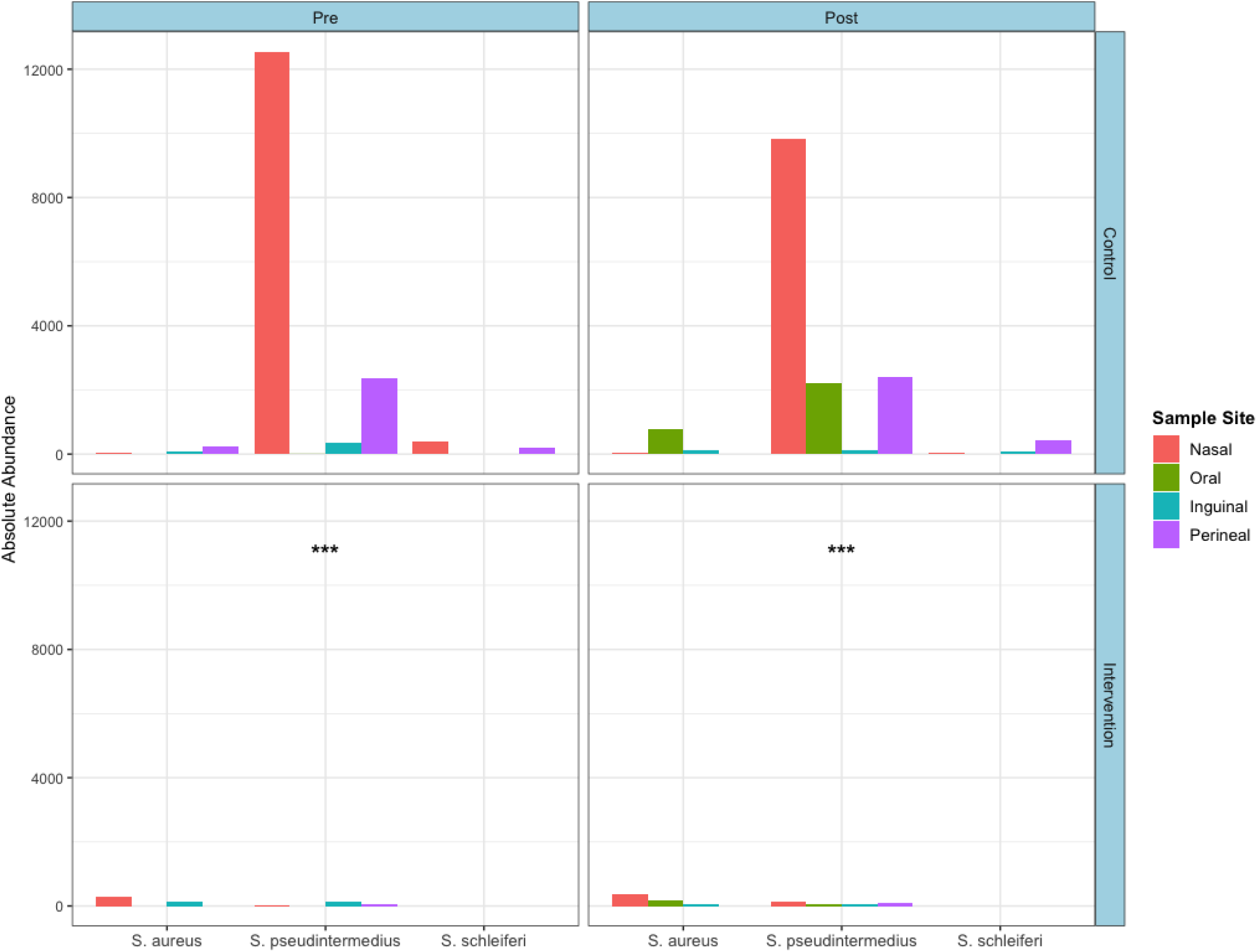
Absolute Abundance of Staphylococcal Species by Site. Bars are aggregated mean absolute abundance of top three *Staphylococcus* species dodged by sample site, stratified by pre and post visit and visit type. *** p<0.0001 DESeq differential absolute abundance by visit type (overall abundance and within nasal samples)

**Supplemental Table 1:**
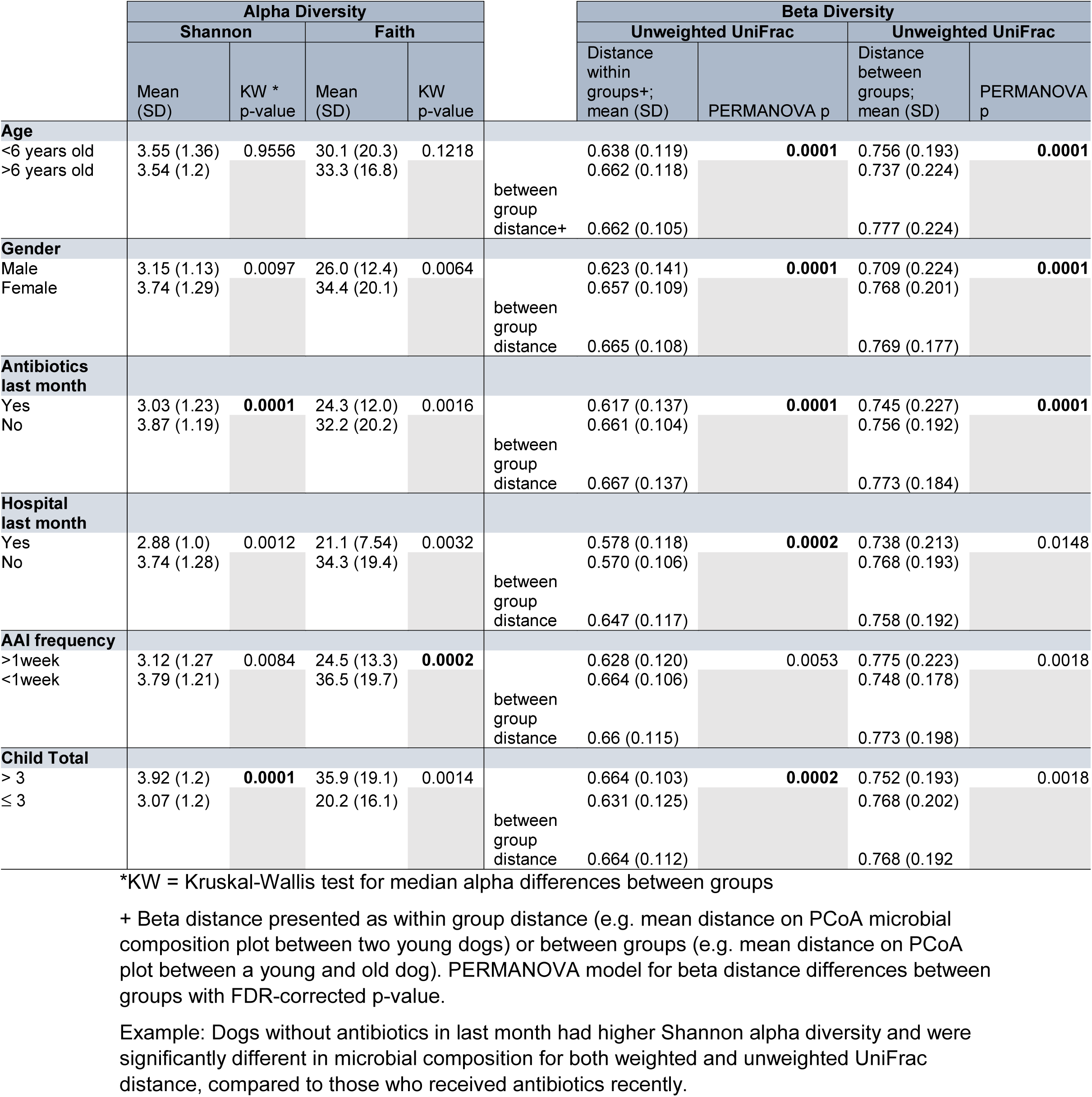
Changes in Alpha and Beta Diversity Levels based on Dog Factors.

